# Suitability of Machine Learning for Atrophy and Fibrosis Development in Neovascular Age-Related Macular Degeneration

**DOI:** 10.1101/2023.03.23.23287652

**Authors:** Jesus de la Fuente, Sara Llorente-González, Patricia Fernandez-Robredo, María Hernandez, Alfredo García-Layana, Idoia Ochoa, Sergio Recalde, Spanish AMD group

## Abstract

Anti-VEGF therapy has reduced the risk of legal blindness on neovascular age-related macular degeneration (nAMD), but still several patients develop fibrosis or atrophy in the long-term. Although recent statistical analyses have associated genetic, clinical and imaging biomarkers with the prognosis of patients with nAMD, no studies on the suitability of machine learning (ML) techniques have been conducted. We perform an extensive analysis on the use of ML to predict fibrosis and atrophy development on nAMD patients at 36 months from start of anti-VEGF treatment, using only data from the first 12 months. We use data collected according to real-world practice, which includes clinical and genetic factors. The ML analysis consistently found ETDRS to be relevant for the prediction of atrophy and fibrosis, confirming previous statistical analyses, while genetic variables did not show statistical relevance. The analysis also reveals that predicting one macular degeneration is a complex task given the available data, obtaining in the best case a balance accuracy of 63% and an AUC of 0.72. The lessons learnt during the development of this work can guide future ML-based prediction tasks within the ophthalmology field and help design the data collection process.

## 1. Introduction

Age-related macular degeneration (AMD) is a progressive chronic disease whose advanced forms, such as neovascular AMD (nAMD), can lead to severe and irreversible vision loss. Neovascular AMD is characterized by macular neovascularization (MNV), which can progress to subretinal fibrosis and macular atrophy (1, 2). Subretinal macular fibrosis is a result of an excessive wound healing response that follows MNV in nAMD and can produce local destruction of photoreceptors, retinal pigment epithelium (RPE) and choroidal vessels (3). On the other hand, macular atrophy is characterized by atrophic lesions of the outer retina, RPE and underlying choriocapillaris, and it is usually found in patients with long-standing nAMD (4). Both atrophy and fibrosis can cause permanent macular dysfunction, legal blindness or inability to perform routine activities such as reading or facial recognition (5).

Advances in diagnostic techniques and anti-vascular endothelial growth factor (anti-VEGF) therapy have helped to reduce AMD-related legal blindness in some countries, and its increasing social and emotional impact (6, 7). However, some patients do not achieve a satisfactory long-term response with current treatment, developing atrophy and fibrosis, and the need for frequent intravitreal injections and ophthalmological visits places a significant burden on patients, their families and healthcare professionals (8).

Some genetic, clinical and imaging biomarkers have been associated with the anatomical and functional prognosis of patients with nAMD, and may help in the planification of individualized anti-VEGF therapies (9–14). One of the imaging biomarkers that has been widely studied in nAMD in the last few years is retinal fluid visualized on optical coherence tomography (OCT), both after the loading phase of anti-VEGF treatment and in the long follow up. The subretinal location of this fluid seems to be related to better visual prognosis and less atrophy and fibrosis formation, while intraretinal fluid has been associated with higher macular fibrosis and worse vision in the long term (13, 15, 16).

The increasing sophistication of imaging systems, networking and software analysis, are making it possible to implement artificial intelligence, such as machine learning (ML), into the diagnostic in medicine, especially in retinal pathologies (17, 18). Nevertheless, in all the aforementioned studies, no ML techniques have been analyzed to predict the outcome of nAMD patients undergoing anti-VEGF treatment. Hence, in this work we evaluate the suitability of ML to predict whether a patient with nAMD will develop fibrosis and/or atrophy after anti-VEGF treatment. We use data collected in a 36-month study according to real-world practice (dataset PI15/01374) to assess possible risk factors in nAMD patients (13). In the previous study, only a conventional statistical analysis of clinical and environmental variables was performed, without evaluation of genetic variables. The objective of this study is therefore twofold: to perform a statistical analysis of the genetic variables that were collected but not analyzed in (13), and to evaluate the predictive power of ML models for atrophy and fibrosis development in nAMD patients at 36 months, using all the clinical and genetic variables collected in routine clinical practice up to 12 months from start of treatment.

## 2. Methods

### 2.1. Study design

Dataset PI15/01374 (13) was used in this study to assess the influence of clinical (including environmental factors) and genetic factors on the progression towards macular atrophy and fibrosis (Table 1 and Supplementary Table S1). Data collection was conducted from 1 September 2016 to 28 February 2020 across 17 sites in Spain, through an ambispective (retrospective and prospective) multicentre 36-month study of a cohort of 354 patients (one eye study) with nAMD treated according to routine clinical practice.

**Table 1.** Feature (variables) contained on the considered dataset PI15/01374. Features are organized in groups based on their clinical similarity.

| Group name | Associated features |  |
| --- | --- | --- |
|  | Numerical | Categorical |
| Atrophy/Fibrosis<sup>†</sup> |  | Atrophy V1, Fibrosis V1, Atrophy V4, Fibrosis V4, Atrophy 12m, Fibrosis 12m, Atrophy 36m, Fibrosis 36m. |
| Demographic | Age | Tabaquism, Sex, Hypertension, Vitamin supplements, Hypercholesterolemia, Dry Macula 36m* |
| Retinal fluid |  | Intraretinal fluid V1, Subretinal fluid V1, Intraretinal fluid V4, Subretinal fluid V4 |
| Foveal thickness | Foveal thickness V1, Foveal thickness V4 |  |
| Neovascular membrane |  | Neovascular membrane V1, Neovascular membrane V4 |
| Cataract |  | Cataract V1, Cataract V4, Cataract 12m |
| ETDRS | ETDRS V1, ETDRS V4, ETDRS 12m |  |
| SNPs |  | ARMS2, CFI, VEGFR, SMAD7, CFB*, CFB1*, CFH*, CFH1*, SERPINF1*, CFI1*, CFI2*, TGFb1*, TNF*, TNF1* |
| Treatment | Injections 36m* |  |

\* These variables have not been included in the ML models due to the lack of importance and improvement within the model or due to data leakage motives.
<sup>†</sup> V1, V4 and 12m variables have not been included in the ML models, see *Data preprocessing* for further details. Atrophy and Fibrosis at 36m are predicting variables.

### 2.2. Genotyping

Genomic DNA was extracted from oral swabs using QIAcube (Qiagen, Hilden, Germany) and processed in the Ophthalmology Experimental Laboratory of the Clínica Universidad de Navarra (Spain). Genetic analysis of 14 single nucleotide polymorphisms (SNPs) was performed by qPCR (Taqman probes): ARMS2 (rs10490924); CFB (rs641153, rs12614); CFH (rs1061170, rs800292); CFI (rs4698775, rs17440077, rs10033900); SERPINF1 (rs12603486); SMAD7 (rs7226855); TGFb1(rs2241713); TNF (rs2256974, rs909253); VEGFR (rs7993418). The SNPs located in the CFB gene were analyzed by Sanger sequencing. The sequence of the Taqman probes for analysis is detailed in Supplementary Table S2. The qPCR was performed with the amplification mix “TaqMan™ Genotyping Mas-ter Mix (Thermo Fisher)” with the specific primers and probes according to the manufacturer’s instructions, in the QuantStudio-5 Applied Biosystem equipment. Controls of known genotype are included in the assay. The analysis of results was carried out with the software: QuantStudio™ Design & Analysis Software. For the genotyping of the SNPs in the CFB gene, the genomic region containing them was amplified with the CertAmp Kit (Biotools) according to the manufacturer’s specifications. The amplification primers are the product of the Secugen design (Forward: 5’ gagccaagcagacaagcaaa 3’(Tm: 61.63ºC); Reverse: 5’ tctccctccccatttctgagt -3’(Tm62.25ºC); Size: 703pb). PCR conditions: 94ºC (3min) +35x [94ºC (0.5min) + 60ºC (1min) + 72ºC (1min)] + 72ºC (10min). The amplicons obtained were visualized on a 2% agarose gel and purified using ExoSAP-IT™ (Applied Biosystems, Spain). Subsequently, they were sequenced by automatic Sanger-type sequencing with BigDye 3.1 reagent and loaded on an AB3730 sequencer. The obtained sequences were analyzed with SeqScape Software v2.5 (Thermo Fisher) (9, 19, 20).

### 2.3. SNPs statistical analysis

To evaluate the significance of the alleles’ frequencies, we used the chi-square test within the following two groups: fibrotic vs non-fibrotic patients, and atrophic vs non-atrophic patients, all at 36 months. All SNPs analyzed in this study were in Hardy-Weinberg equilibrium. The Bonferroni method was used to correct for multiple comparisons. The results of this analysis are also used to perform feature selection of the genetic variables prior to the ML model (see Data preprocessing subsection).

### 2.4. Machine Learning analysis

The dataset PI15/01374 specifies whether a nAMD patient developed fibrosis and/or atrophy at 36 months. Due to the different nature of these outcomes, we considered distinct machine learning models to predict, at 12 months from start of treatment, whether a patient (eye) will develop 24 months later (i.e., at 36 months): atrophy and/or fibrosis (Atrophy|Fibrosis_36m); fibrosis (Fibrosis_36m); and atrophy (Atrophy_36m). In all cases this reduces to a supervised learning problem for binary classification, in which the positive class is referred as having the pathology and the negative class as not having it.

#### 2.4.1. Data preprocessing

The considered PI15/01374 dataset contains information of clinical and genetic (SNPs) variables for 335 eyes. Before being used as input to the ML models, we performed some preprocessing steps.

Since the goal is to make a prediction on month 12 after starting the treatment, clinical variables collected at 36 months were removed, as they would not be available in a predicting real scenario. This reduced the number of clinical variables to 20 (see Table 1). Out of the 14 genetic variables, we selected a representative SNP from each of the 4 risk-pathways associated with nAMD Atrophy and Fibrosis: complement system (*CFI*), metabolic change in mitochondria (*ARMS2*), inflammation (*SMAD7*) and neovascularization (*VEGFR*) (21). The SNPs statistical analysis results were used to guide this selection and filter out SNPs that did not show statistical differences (Figure 1). To ease the feature importance analysis (see subsection 2.4.4), the retained clinical and genetic variables were further split in seven groups based on their clinical similarity (Table 1). Due to the high variables/eyes ratio, categorical variables (including SNPs) were encoded following a LabelEncoding instead of a OneHotEncoding (using Python’s sklearn library).

**Fig. 1.**
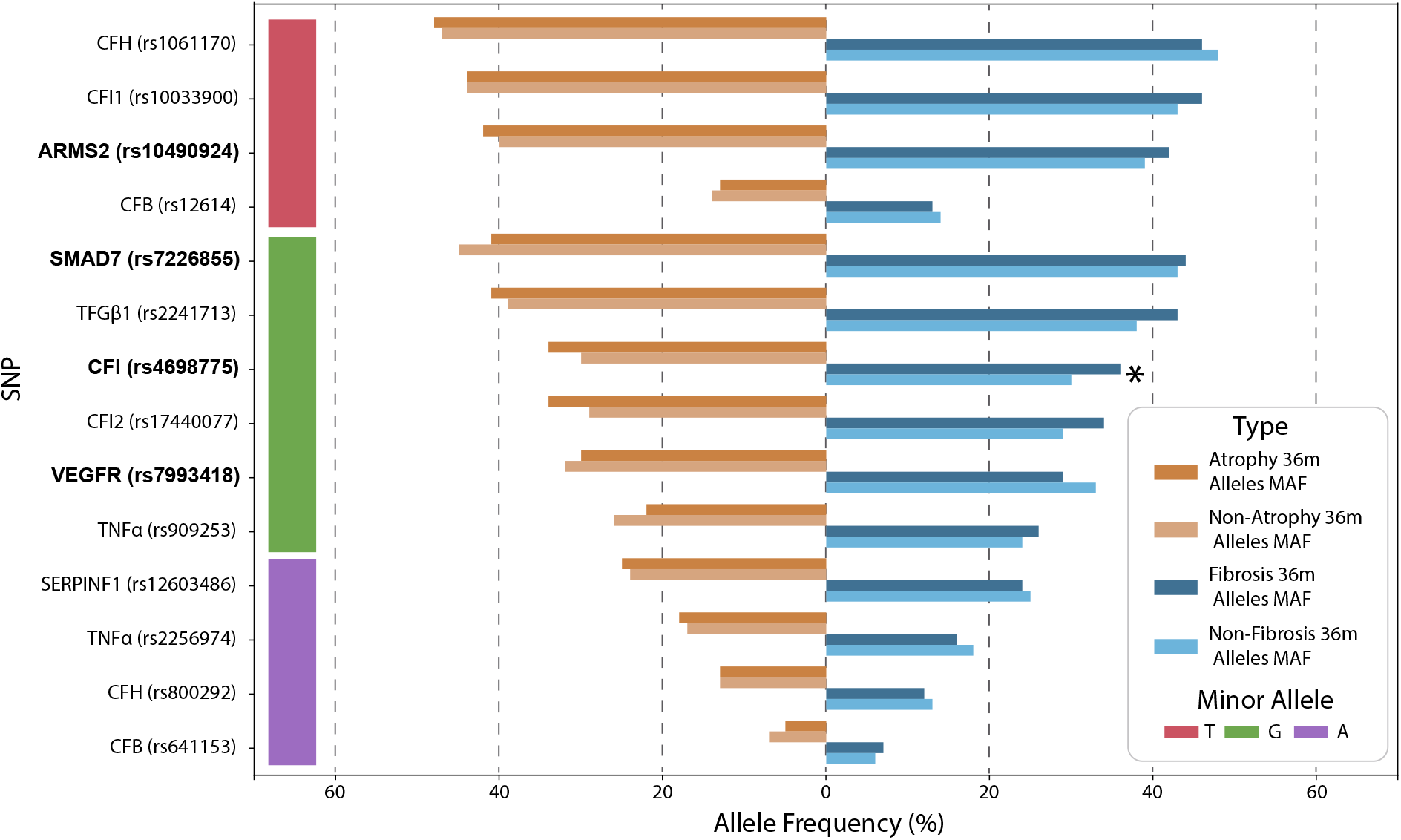
Minor Allele Frequency (MAF) differences between Atrophy/non-Atrophy and Fibrosis/non-Fibrosis patients. Two barplots have been used for representing the MAF alleles frequencies for each SNP and disease of study. Minor allele type for each SNP has also been added. The significant allele frequency differences, according to the chi-square test among groups within the same disease (see Methods), have also been pointed out (*). SNPs used in the ML models (see Table 1) are highlighted in bold.

We dropped samples (eyes) which already presented the pathology to be predicted at 4 or 12 months, as it was observed that in these cases the pathology remained unchanged at 36 months. Moreover, retaining these samples can oversimplify the models and avoid their correct training. We also dropped samples containing variables (from the retained ones) with missing values (N/A). These steps reduced down the number of samples to 296 for the atrophy experiment, 284 for fibrosis and 254 for atrophy and/or fibrosis. In total, 55% of the samples presented atrophy and/or fibrosis at 36 months, 37% presented fibrosis, and 30% presented atrophy.

#### 2.4.2. Supervised learning models

Three different supervised learning methods known to perform well in practice were selected: Random Forest (RF) (22), Extreme Gradient Boosting (XGB) (23) and Support Vector Machines (SVM) (24). Deep learning models were not considered due to the low number of available samples. RF and XGB are encompassed within the field of ensemble learning, as they combine decision trees (DTs) to find patterns and classify the data. RF is based on bagging, which performs bootstrapping over the data and uses multiple DTs to average the results and reduce the variance. To decorrelate the trees and prevent overfitting, in RF the DTs can only use a random subset of the features. XGB is based on boosting, in which trees are built sequentially (i.e., previously built trees are taken into account to build the next one). SVM classifies the data by applying linear separators, making use of kernels to get margin classifiers that work efficiently in very high dimensional data. Both RF and XGB fall within the category of soft-classifiers, as they compute the posterior probability of an input sample belonging to the positive class. SVM is a hard-classifier that outputs the predictive class without explicitly computing the posterior probability. Yet, an estimation of this probability can be computed using cross-validation. By default, if the posterior (or predictive) probability is larger or equal to 0.5, a positive prediction is made (negative otherwise). Nevertheless, since these probabilities reflect how confident the model is when making a prediction, a different threshold (Th) can be used such that only samples with a probability greater than Th are classified as positive. As shown below, the capacity of a model to separate both classes can be evaluated by modifying this threshold.

#### 2.4.3. Evaluation metrics

Accuracy, defined as the percentage of samples correctly classified (i.e. for which the correct prediction is made), is generally the preferred metric to evaluate ML models for classification. However, due to the data imbalance among positive and negative samples, balanced accuracy (BA) score was also considered. BA computes the average between the accuracy on the positive samples and the accuracy on the negative samples, giving equal weight to both classes. To evaluate the reliability and confidence of the models, we considered the Area Under the ROC (Receiver Operating Characteristic) Curve (AUC). The ROC curve plots the True Positive Rate (TPR) vs the False Positive Rate (FPR) for each possible threshold, defined as:

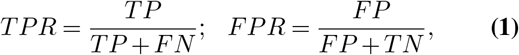

where TP, FN, and FP stand for true positives, false negatives, and false positives, respectively. The AUC is given by the area under the ROC curve, and ranges from 0 to 1, with 0.5 being a random classifier and 1 a perfect one. Intuitively, a reliable and confidence model should generate high probabilities when input positive samples, and viceversa. Additionally, if samples are sorted by their predictive probabilities, positive samples are expected to appear before negative samples, such that for high thresholds only positive samples would be predicted as positive (i.e., FPs would be close to zero). As the threshold decreases, the opposite is expected, i.e., we should have close to zero FNs. The ROC curve and the AUC therefore provide metrics to better understand how well the model separates both classes.

#### 2.4.4. Feature importance

In order to analyze the models’ feature importance in a homogeneous manner, we define a relative AUC (rAUC) score as

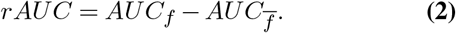

rAUC measures the increase or decrease in the models’ AUC that a specific feature or group of features yield. *AUC*_*f*_ accounts for the AUC of a model when using a group of features as input, including the specific feature *f* we want to compute the rAUC for. 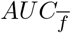 accounts for the AUC of the same model, i.e., same parameters and same features, but excluding feature *f*. Hence, rAUC measures the specific contribution of feature *f* to the model’s reliability. For a set of *p* features, there would be 2^*p*^ − 1 possible combinations. Hence, to reduce the computation complexity, we analyze the importance of each feature group (Table 1) rather than individual features, and apply this metric to models with at least two group of features. 2^*p*^ − 1 − *p* combinations are therefore evaluated (*p* being 7 in our case).

#### 2.4.5. Experimental setup

When evaluating ML models, it is key to verify their generalization ability, i.e., how they perform on data not used for training (referred to as test data). Due to the low number of available samples, cross-validation (CV) was used to generate training and test folds iteratively (25). However, when performing hyperparameter tuning and feature selection simultaneously, CV can yield overfitted test folds. In our case, for each ML model, we considered different values for the hyperparameters as well as all combinations of feature groups. Therefore, we used nested cross validation (NCV) instead (26). Similarly to CV, in NCV data is split in folds, and at each iteration one fold is left out for testing and the remaining ones are used for training (called outer training fold in NCV). But contrary to CV, in NCV the outer training fold is further split into folds, and iteratively all folds but one are used for training and the left out fold for validation. The hyperparameters that better perform (on average) in the validation sets are then tested in the test fold. This allows hyperparameter tuning and feature selection while ensuring generalization ability of the resulting models, avoiding overfitting and increasing robustness during the training.

In our experiments 6 folds were used in both the outer and inner loops. For each considered model, hyperparameter tuning was performed by applying a grid search (Supplemen-tary Table 4), and all possible subsets of the defined feature groups were tested during training. For each prediction task, we evaluated the importance of each feature group by computing the corresponding rAUCs on the test sets from NCV. Finally, the model with the best combination of features and hyperparameters in terms of average AUC (on the test folds) was selected. Unless stated otherwise, all reported metrics are on the test folds (from NCV).

## 3. RESULTS

### 3.1. SNPs statistical analysis results

The results of the allelic analysis of the 14 considered SNPs regarding their association with the development of atrophy or fibrosis are shown in Figure 1. Allelic frequencies exhibited a significant association between patients with fibrosis compared to non-fibrotic patients with the CFI gene. All the SNPs of this gene showed frequencies with some differences (Supplementary Table S3) but the SNP rs4698775 (CFI) indicated a significantly higher minor allele frequency (MAF) in patients with fibrosis (p<0.05, OR 1.4 with 95% CI 1.0-1.9) versus non-fibrotic. This significance is however lost after Bonferroni adjustment (p>0.05). Regarding the development of atrophy, no significant differences were found in the allelic frequencies of these 14 SNPs.

### 3.2. Machine learning results

#### 3.2.1. Feature importance

We first evaluated the importance of each of the seven considered feature groups by computing the corresponding rAUC values. Due to the flexibility of RF, XGB and SVM models and the limited availability of samples, we considered different hyperparameters for each model and prediction task, as well as all possible combinations of feature groups. Figure 2 shows the rAUC distributions across features group and ML models, for each prediction task.

**Fig. 2.**
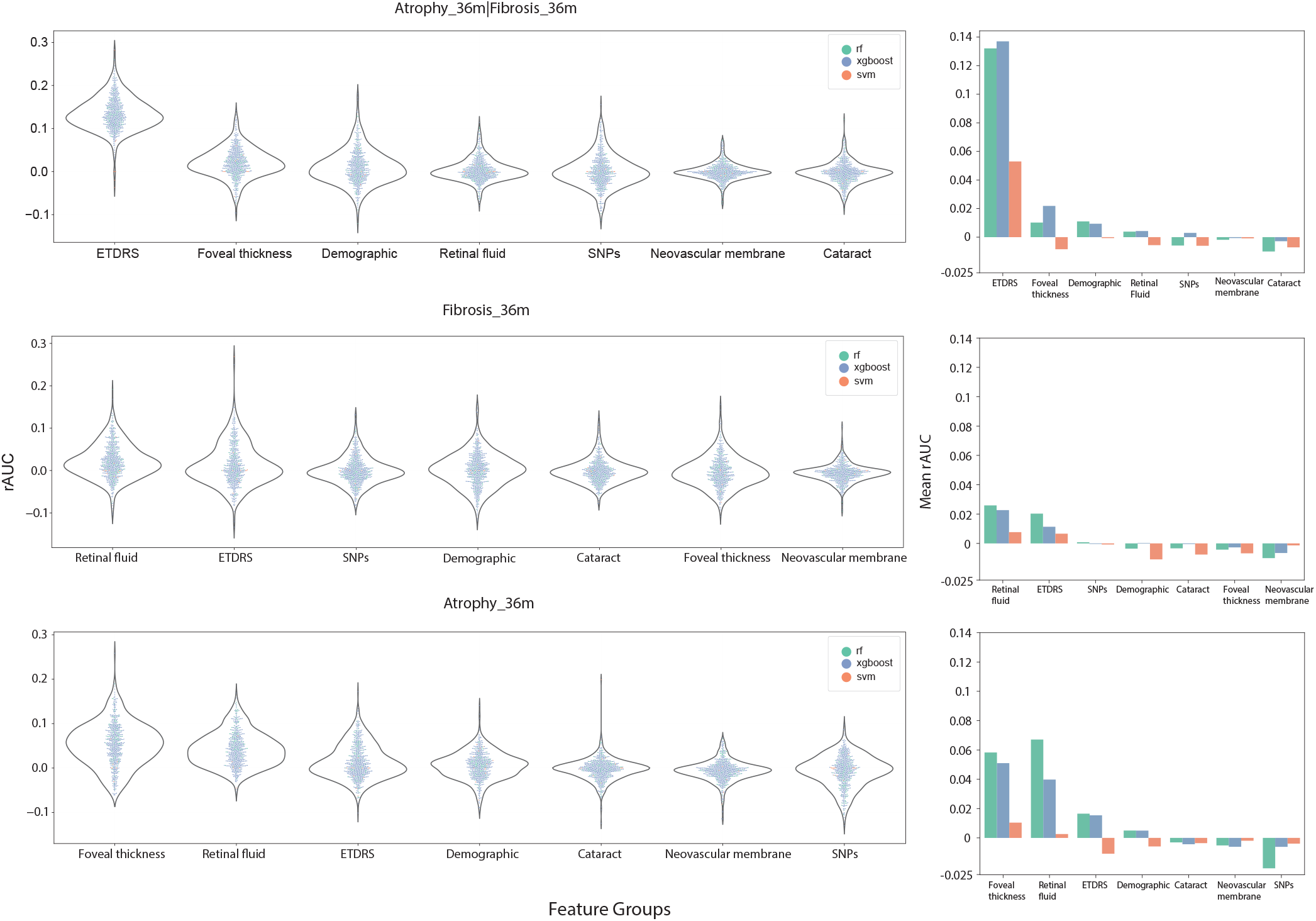
Relative AUC (rAUC) scores for each group of features and prediction task. Left. Distribution of rAUC values as a function of each group of features, for the three prediction tasks, shown as a violin plot. A swarm plot is added within each violin plot to distinguish among the three machine learning models (RF, XGB, SVM). **Right**. Barplot showing the mean rAUC as a function of each group of features, for every prediction task.

##### Atrophy or Fibrosis at 36m

When analyzing the Atrophy|Fibrosis_36m experiment, we observed that ETDRS and foveal thickness were the two most important feature groups based on rAUC, especially for the XGB model (Figure 2). The importance of ETDRS is related to the previous statistical analysis, where it was shown that atrophy and fibrosis at 36 months were associated with lower ETDRS at any visit, explained by the visual impairment generated at the macular level (13, 15). However, foveal thickness at baseline (V1) and after the loading phase (V4) did not show statistically significant differences for the development of atrophy and fibrosis in the previous statistical analysis (p>0.05). This is not surprising, as ML models can learn complex patterns in the data, and features that are not statistically significant when analyzed in isolation may add relevant information to the models when combined with other features. The fact that all features in the ETDRS and foveal thickness groups are numerical may also help, as numerical variables can ease the exploitation of patterns in ML models with high predictive power such as the ones being evaluated. Interestingly, even though the variance of rAUC for ETDRS variables is larger than the variance of rAUC for foveal thickness variables, the ETDRS rAUC score distribution is significantly higher than the one of foveal thickness (p<0.001), corroborating the importance of ETDRS in the evolution of nAMD. It is worth noting that contrary to what previous studies have shown related to the statistical power of the retinal fluid variable group to predict atrophy and fibrosis diseases in nAMD patients, their rAUC distribution shows that they do not add much value to the ML models. Probably, the fact that the retinal fluid variables are qualitative has caused them to be less relevant for the predictive models, while foveal thickness (quantitative), being directly correlated with retinal fluid (since the greater the fluid, the greater the foveal thickness and vice versa), would be an indirect reflection of the importance of retinal fluid. Future studies should consider collecting the retinal fluid variables as quantitative rather than qualitative. Finally, demographic, cataract, SNPs and neovascular membrane groups do not seem to add value to the ML models in terms of rAUC (distribution centered around 0).

##### Fibrosis at 36m

In the Fibrosis_36m prediction task, the ETDRS rAUC distribution shows a similar but much less pronounced trend to that of the Atrophy|Fibrosis_36m experiment. ETDRS and retinal fluid are the only two groups with a non-negative mean rAUC (see Figure 2). The fact that the rAUC distribution is more skewed towards smaller values as compared to the Atrophy|Fibrosis_36m rAUC distribution can be associated with the complexity of the predicted variable. Specifically, in the Fibrosis_36m experiment, both healthy patients and those that develop only atrophy at 36 months belong to the negative class, even though the latter also have a bad prognosis. This can add noise and blur the decision making of the ML models. On the contrary, Atrophy|Fibrosis_36m includes patients that develop either atrophy or fibrosis in the positive class, avoiding this problem.

##### Atrophy at 36m

Finally, the rAUC distributions in the Atrophy_36m experiment show foveal thickness and retinal fluid groups to increase the robustness of the model the most (see Figure 2). This can be explained by the relation between these groups of variables, as mentioned above, and the fact that retinal fluid has been previously identified as having clinical importance in the development of atrophy in nAMD (13, 15).

#### 3.2.1. Predictive results

##### Model selection

After the conducted analysis that considered all combinations (>250,000) of ML models (RF, XGB and SVM), hyperparameters and feature groups, the combination with the highest validation AUC score (during NCV) was selected for each prediction task. Table 2 contains a summary of the final models. In all cases XGB obtained the highest AUC, albeit with a different set of hyperparameters. Regarding the feature groups, all three experiments employ the ETDRS group and an additional feature group. Specifically, the Atrophy|Fibrosis_36m experiment includes the foveal thickness group, the Fibrosis_36m experiment the SNPs, and the Atrophy_36m experiment the retinal fluid group.

**Table 2.** Detailed information about the final models used for each prediction task, including hyperparameters and input features.

| Experiment | Atrophy\|Fibrosis_36m | Fibrosis_36m | Atrophy_36m |
| --- | --- | --- | --- |
| Model | XGBoost | XGBoost | XGBoost |
| Variables | ETDRS<br>Foveal Thickness | ETDRS<br>SNPs | ETDRS<br>Retinal fluid |
| Hyperparameters |  |  |  |
| Number of estimators | 287 | 287 | 525 |
| Max depth | 50 | 50 | 50 |
| Min child weight | 0.1 | 5 | 0.1 |
| Gamma | 1.5 | 0.5 | 1.5 |
| Subsample | 1.0 | 1.0 | 1.0 |
| Colsample by tree | 0.6 | 0.6 | 1.0 |
| Learning rate | 0.0001 | 0.1 | 0.0001 |
| Reg alpha | 0.1 | 0.0001 | 0.1 |
| Reg lambda | 0.0001 | 0.1 | 0.0001 |

##### Performance metrics

Next we report the obtained evaluation metrics of the final models for each prediction task, computed as the average across the NCV test folds (see Figure 3A). For Atrophy|Fibrosis_36m, the obtained average BA is 0.63, the accuracy 0.65, and the AUC 0.72. For Fibrosis_36m, the average BA is 0.54, the accuracy is 0.72, and the AUC is 0.6. Finally, for Atrophy_36m, the average BA is 0.54, the accuracy is 0.7, and AUC is 0.57. Due to the imbalance between positive and negative samples in all experiments, BA is always lower than accuracy, showcasing the importance of considering BA in addition to accuracy. The highest AUC is obtained for the Atrophy|Fibrosis_36m, since there is a more clear distinction between negative and positive samples.

**Fig. 3.**
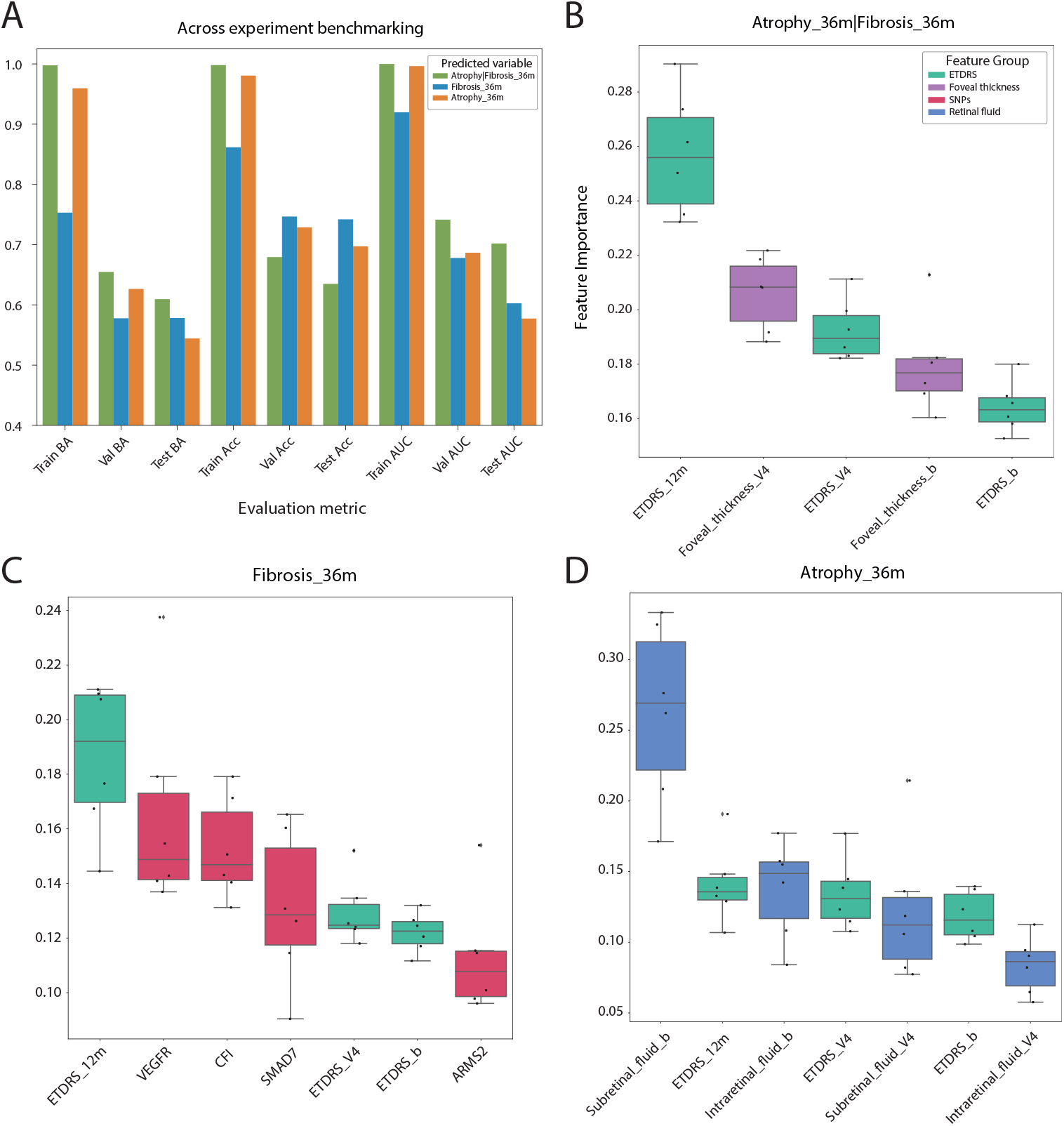
Evaluation metrics and importance scores of the final models for each prediction task. A. Evaluated metrics (BA, accuracy and AUC) along the folds (train, validation and test) for each experiment for the final ML models. **B-D**. Boxplots showing the distribution of importance scores for variables within each group from the test split across folds of the NCV setup, for **B)** Atrophy|Fibrosis_36m, **C)** Fibrosis_36m, and **D)** Atrophy_36m experiments. Variables within each experiment are sorted by their corresponding feature importance mean.

To further assess the proposed models, Figure 3A shows the evaluation metrics obtained for each of the splits within NCV. It is clear from the results that there is an intrinsic complexity in the prediction of atrophy and fibrosis given the available data. This is more pronounced for the Fibrosis_36m and Atrophy_36m experiments, in which lower metrics are obtained as compared to the Atrophy|Fibrosis_36m experiment. As stated above, this is expected, as the prediction task in the first two experiments is more complex. Moreover, results for Atrophy|Fibrosis_36m and Atrophy_36m show signs of overfitting in the training fold, suggesting the models were complex enough to learn complex patterns, but the variance within patients did not allow these patterns to become gener-alizable. The same trend is found in the Fibrosis_36m experiment. Even though the accuracy does not show overfitting signs, patients from validation and test folds yield lower accuracies and AUCs, highlighting the underlying difficulty of the prediction tasks. This is reasonable, since heterogeneity has been pointed out as a common denominator in patients with nAMD, and more so in this real-life clinical practice study, with less exhaustive inclusion and exclusion criteria than in a clinical trial, applying various anti-VEGF therapies, multiple treatment and follow-up regimens.

##### Feature importance

We also analyzed the importance of each of the included features, computed as the average gain across all splits the feature is used in (gain option in the xgboost Python package).

###### Atrophy or Fibrosis at 36m

Regarding the Atrophy|Fibrosis_36m experiment, even though ETDRS at V1 (ETDRS_b) and 4 months (ETDRS_V4) do not show relevance signs within the best combination, ETDRS at 12 months is statistically significant (from the feature importance perspective) for the classification power of the model (p<0.001, Figure 3A). The rationale behind this is that ETDRS variables are correlated within each other (Supplementary Figure S1) and the model uses only one (the closest in correlation to the predicting variable) for most of the splits, hence obtaining the highest feature importance among the three ETDRS features and in general also.

###### Fibrosis at 36m

The Fibrosis_36m prediction task exhibits a similar feature importance distribution to the Atrophy_36m|Fibrosis_36m experiment (Figure 3B). The importance of ETDRS at 12 months is also significantly above the rest of variables (p<0.05), followed by the SNPs VEGFR, CFI and SMAD7. Recall that the SNP CFI showed some statistical differences between fibrotic and non-fibrotic patients. Finally, as expected, the order importance of the ETDRS variables are sorted by time (12 months, V4, and V1).

###### Atrophy at 36m

Finally, for the Atrophy_36m experiment, the importance of the basal subretinal fluid appears to be significantly above the other features (p<0.001, Figure 3C). Clinically, subretinal fluid has shown to be associated with a better visual acuity and a lower risk of developing macular atrophy or fibrosis, with fewer injections (11–13). The remaining features do not seem to add to the predictive power of the model. Nevertheless, the performance of the model (BA 0.54 and AUC 0.57) indicates that the model cannot learn to distinguish between atrophic and non-atrophic patients. This is expected, as the development and evolution of atrophy involves the interaction of several metabolic, functional, genetic and environmental factors, making its affectation unpredictable (27). Likewise, at a functional level, atrophy can appear in an advanced form but not have much visual affectation and vice versa, making its prediction very complex.

## 4. DISCUSSION AND CONCLUSION

This work presents, to the best of our knowledge, the first exhaustive analysis regarding the suitability of machine learning for predicting development of fibrosis and atrophy on neovascular age-related macular degeneration patients undergoing anti-VEGF treatment. The ML models are trained to predict the development of fibrosis and atrophy at 36 months after starting the treatment with VGEF, using data collected during the first 12 months. For the analysis, we used demographic, clinical, and genetic variables. We consistently found ETDRS to be relevant for the prediction of atrophy and fibrosis, confirming previous statistical analyses (13). On the other hand, the analyzed SNPs, being in some cases widely associated with AMD development (with high risk or protective frequencies compared to healthy controls), have not shown any specific association with macular degeneration in the considered cohort, and have not significantly contributed to the ML models. The best performing model is able to predict the development of at least one macular degeneration with an accuracy of 65%, a balance accuracy of 63%, and an AUC of 0.72. As highlighted below, access to more samples as well as more features (or of better quality) could boost the prediction power of ML models.

In particular, even though the presented results confirmed the known relationship between macular degeneration and retinal fluids on OCT (11–13, 28, 29), we believe that the categorical nature of these features may have narrowed down the pattern-exploitation ability of the applied ML predictors. Hence, storing the numerical value (OCT liquid volume) for these features may help in future ML studies.

Moreover, due to the nAMD heterogeneity and the underlying complexity of atrophy and fibrosis diseases, the ML models would benefit from a higher number of samples, as the lack of generalization has been observed along the three considered experiments. Still unexplored deep learning approaches would also benefit from this.

Regarding the evaluated SNPs, even though they did not show to be sufficient to predict nAMD development, additional analysis with larger cohort of patients should be carried out before they are ruled out, as they could have a regulatory role in these processes. A different set of SNPs could also be evaluated to analyze their potential effect on disease progression.

The fact that nAMD is a complex disease involving many factors means that the ML models need access to high quality data in order to make accurate predictions. Hence, when collecting data from real clinical practice, it would be desirable to use the same (or similar) image detection and analysis systems, so that data is as homogeneous as possible, and to have long follow-up periods with regular visits, so that more information per patient is available. Raw values should also be collected for each variable when possible, e.g., without converting numerical variables to categorical by applying thresholds.

In summary, in this work we have established the guidelines for future nAMD atrophy and fibrosis prediction. Several ML approaches have been analyzed and, despite the complexity of the prediction task, multiple already-known biological relationships have been found along the process. Moreover, lessons learnt during the development of this work may guide future ML-based prediction tasks within the ophthalmological field and help design the data collection process.

## Data Availability

All data produced are available online at the provided github repository.

https://github.com/jesusdfc/ml_namd

## Code and Data availability

All supporting code, data (PI15/01374) and materials are available in the following github repository https://github.com/jesusdfc/ml_namd.

### List of abbreviations

Anti-VEGF: Anti–Vascular Endothelial Growth Factor
nAMD: neovascular Age-related Macular Degeneration
MAF: minor allele frequency
SNP: Single Nucleotide Poly-morphisms
ETDRS: Early Treatment Diabetic Retinopathy Study
OCT: Optical coherence tomography
V1: baseline visit
V4: after the loading phase treatment visit
MNV: macular neovascularization
SRF: subretinal fluid
IRF: intraretinal fluid
RPE: retinal pigment epithelium
ML: Machine Learning
XGB: eXtreme Gradient Boosting
RF: Random Forest
SVM: Support Vector Machine
NCV: Nested Cross-Validation
BA: Balanced Accuracy
AUC: Area under the Curve
rAUC: relative AUC
OD: Odds ratio
CI: Confidence interval
ROC: Receiver Operating Characteristic
CV: Cross-Validation
DT: Decision Tree
Th: Threshold
TPR: True Positive Rate
FPR: False Positive Rate
TP: True Positive
FN: False Negative
FP: False Positive
ROC: Receiver Operating Characteristics

## Ethical Approval and consent to participate

All procedures carried out in this study were in accordance with the guidelines of the Declaration of Helsinki. The Institutional Review Board and the Ethics Committee of Clinica Universidad de Navarra (CUN-RAN-2016-01) and Government of Navarra, Spain (EO16/19), approved the protocols used in this study. All patients were fully informed of the purpose and procedures, and written consent was obtained from each patient.

## Author’s Contributions

J.F: ML model and analysis, writing, editing, data uploading. S.L.G: Data uploading, experiment, database, writing, and editing. PFR: Study Design, database and editing MHS:database and editing. AGL: Study Design, Data uploading, reviewing and supervision. I.O: writing, reviewing and supervision. SR: Study Design, database, writing, reviewing and supervision. Spanish AMD Group: Data uploading. All authors contributed to the article and approved the submitted version.

## Supplementary Materials

**Supplementary Table S1**. Distribution of atrophy and fibrosis at 36 months according to demographic and clinical characteristics.

**Supplementary Table S2**. Taqman sequences for SNPs. **Supplementary Table S3**. Minor Allele Frequency differences.

**Supplementary Table S4**. Hyperparameter tuning parameters.

**Supplementary Figure S1**. Spearman correlation between features computed across all available eyes.

