## Supplementary figures and images for "Suitability of Machine Learning for Atrophy and Fibrosis Development in Neovascular Age-Related Macular Degeneration"

### Supplementary Figure 1

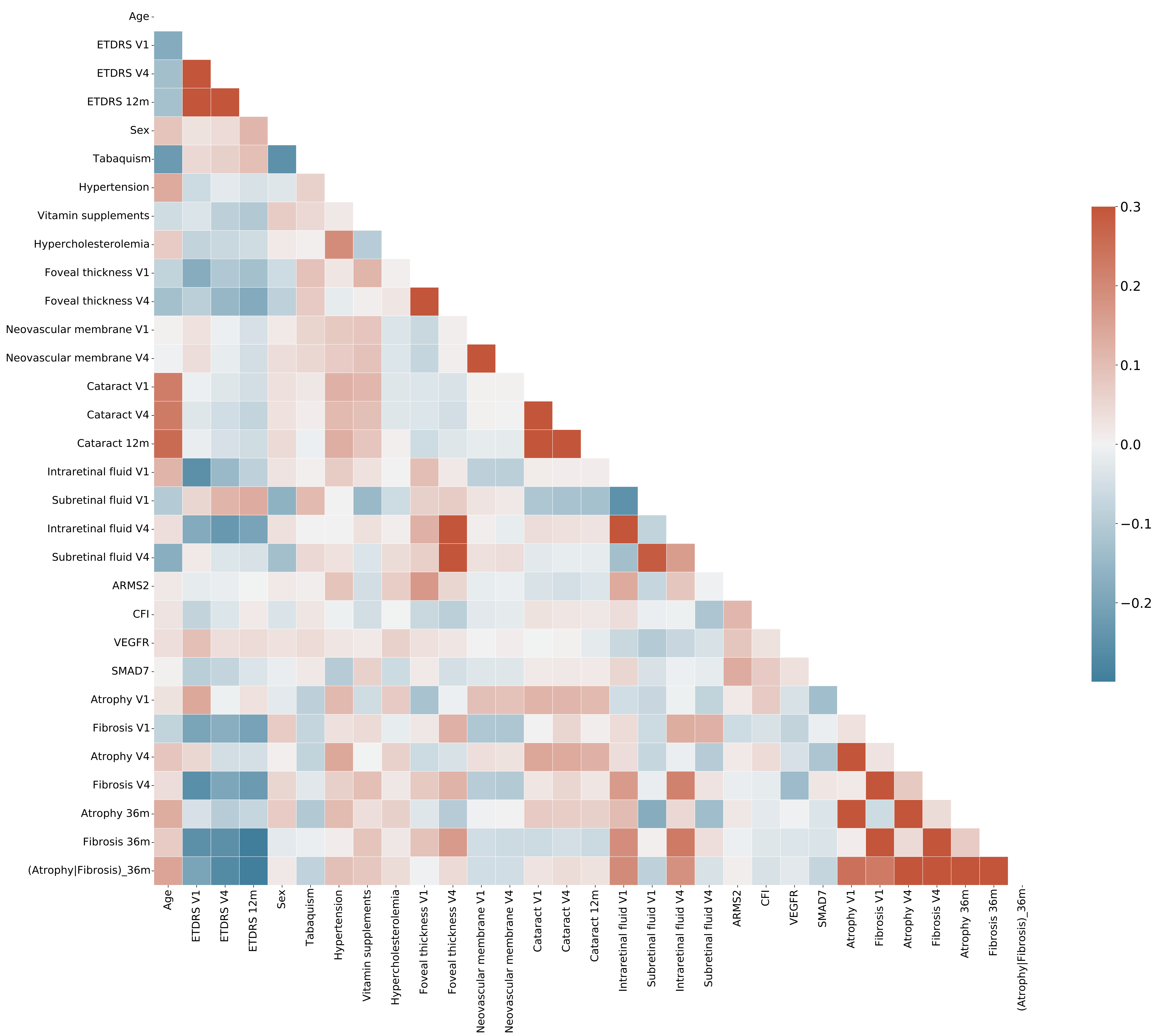
