## Supplementary Table 1 for "Suitability of Machine Learning for Atrophy and Fibrosis Development in Neovascular Age-Related Macular Degeneration"

**Supplementary Table S1. Distribution of atrophy and fibrosis at 36 months according to demographic and clinical characteristics.** V1: baseline visit; V4: visit after loading phase; 12: 12-month visit; 36: 36-month visit; MNV 1: macular neovascularization type 1; MNV 2: macular neovascularization type 2; mixed: combination of MNV type 1 and 2; SRF: subretinal fluid; IRF: intraretinal fluid; N: number of patients; A: atrophy; F: fibrosis; OR: odds ratio; 95% CI: 95% confidence interval. T student was performed to evaluate statistically significant differences (p<0.05).

| | **Mean ± SD**<br>**N=354 (%)** | **A 36**<br>**N=111**<br>**(31.3 %)** | **No A 36**<br>**N=238**<br>**(67.2%)** | ***OR***<br>***(95% CI)***<br>**P** | **F 36**<br>**N=134**<br>**(37.8%)** | **No F 36**<br>**N=215**<br>**(60.7%)** | ***OR***<br>***95% CI***<br>**P** |
|---|---|---|---|---|---|---|---|
| **Age (years)** | 76.7±7.1 | **78.0±6.5** | 76.1±7.2 | **0.023** | 77.3±6.3 | 76.4±7.4 | 0.22 |
| **Female sex** | 215 (60.7) | 74 (66.7) | 138 (57.9) | *OR 1.4*<br>*(0.9-2.5)*<br>0.12 | 80 (59.7) | 132 (61.4) | *OR 1.0*<br>*(0.6-1.6)* 0.82 |
| **Arterial hypertension** | 223 (63) | 76 (68.5) | 144 (60.5) | *OR 1.4*<br>*(0.9-2.5)*<br>0.15 | 83 (61.9) | 137 (63.7) | *OR 1.0*<br>*(0.7-1.7)* 0.81 |
| **Hypercholesterolemia** | 150 (42.4) | 51 (45.9) | 97 (40.7) | *OR 1.25*<br>*(0.7-2)*<br>0.42 | 58 (43.3) | 90 (41.8) | *OR 1.1*<br>*(0.7-1.7)*<br>0.82 |
| **Smoking** | 77 (21.7) | **16 (14.4)** | 60 (25.2) | ***OR 0.5***<br>***(0.3-0.9)***<br>**0.025** | 27 (20.1) | 49 (22.8) | *OR 0.9*<br>*(0.5-1.4)*<br>0.59 |
| **ETDRS V1 (letters)** | 57.1±16.8 | 55.5±18.3 | 57.6±16.6 | 0.27 | **48.7±19.7** | 60.5±14.7 | **<0.001** |
| **Foveal thick. V1 (micron)** | 328.7±89.2 | 348.3±85.3 | 333.4±79.5 | 0.895 | 346.3±92.8 | 323.7±87.7 | 0.099 |
| **MNV V1: 1**<br>**2**<br>**Mixed** | 203 (57.3)<br>96 (27.1)<br>55 (15.5) | 58 (52.2)<br>26 (23.4)<br>**27 (24.3)** | 141 (59.2)<br>70 (29.4)<br>27 (11.3) | ***OR 2.4***<br>***(1.3-4.5)***<br>**0.005** | 72 (53.7)<br>44 (32.8)<br>18 (13.4) | 127 (59.1)<br>52 (24.2)<br>36 (16.7) | *OR 1.5*<br>*(0.9-2.4)*<br>0.12 |
| **SRF V1** | 279 (79.3) | 75 (68.1) | **201 (84.4)** | ***OR 0.4***<br>***(0.2-0.7)***<br>**<0.001** | 106 (79.1) | 170 (79.1) | *OR 0.9*<br>*(0.6-1.6)*<br>1.0 |
| **IRF V1** | 214 (61) | **74 (67.2)** | 138 (57.9) | ***OR 1.87 (1.1-3.2)***<br>**0.021** | **96 (71.6)** | 116 (53.9) | ***OR 2.2***<br>***(1.4-3.6)***<br>**0.001** |
| **ETDRS V4 (letters)** | 64.6±14.5 | 62.6±14.2 | 65.4±14.7 | 0.09 | **59.8±16.8** | 67.4±12.2 | **<0.0001** |
| **SRF V4** | 125 (35.3) | 30 (27.5) | **95 (40.0)** | ***OR 0.6***<br>***(0.3-0.9)***<br>**0.03** | 52 (39.3) | 73 (33.9) | *OR 1.2*<br>*(0.8-2.0)*<br>0.35 |
| **IRF V4** | 93 (26.3) | 34 (31.2) | 59 (24.7) | *OR 1.4*<br>*(0.8-2.3)*<br>0.24 | **53 (40.1)** | 40 (18.6) | ***OR 2.9***<br>***(1.8-4.8)***<br>**<0.0001** |
| **ETDRS 12 (letters)** | 63.3±16.6 | 61.3±17.2 | 64.0±16.4 | 0.16 | **56.1±19.9** | 67.5±12.4 | **<0.0001** |
| **ETDRS 36 (letters)** | 58.5±20.7 | 56.1±20.5 | 59.5±20.8 | 0.17 | **48.3±24.0** | 64.7±15.5 | **<0.001** |
| **Injections 36** | 13.8±5.3 | 13.1±4.5 | 14.3±5.4 | 0.56 | 13.1±4.7 | 14.5±5.4 | 0.42 |
