## Supplementary Table 2 for "Suitability of Machine Learning for Atrophy and Fibrosis Development in Neovascular Age-Related Macular Degeneration"

**Supplementary Table S2: Taqman sequences for SNPs.** The sequence of the Taqman probes for each analyzed SNP except the SNPs located in the CFB gene (rs641153, rs12614) that were analyzed by Sanger sequencing.

| Gen | SNP ID | Label | Sequences |
|---|---|---|---|
| *ARMS2* | rs10490924 | [Vic/Fam] | TATCACACTCCATGATCCCAGCT[G/T]CTAAAATCCACACTGAGCTCT |
| *CFH* | rs1061170 | [Vic/Fam] | AAAATGGATATAATCAAAAT[C/T]ATGGAAGAAAGTTTGTACAG |
| *CFH* | rs800292 | [Vic/Fam] | TGGATATAGATCTCTTGAAAT[A/G]TAATAATGGTATGCAGGAAGGGA |
| *CFI* | rs4698775 | [Vic/Fam] | TTAGGCTGCTTTGTTTTTTTCTC[G/T]GAATGCTAAATATTTTATCCCA |
| *CFI* | rs17440077 | [Vic/Fam] | CACAGTACCCTACCTCTAGTGGA[A/G]TATGCACAATAGGGCTGTATT |
| *CFI* | rs10033900 | [Vic/Fam] | TCAGGTCATCTCACTCCTGCTGT[C/T]CCTGGTCTCTGTCACATAGAG |
| *SERPINF1* | rs12603486 | [Vic/Fam] | GATGGTGAGGCCTCTGTCTTGC[A/G]CTGCAGAAAGCTTTTCCTGTT |
| *SMAD7* | rs7226855 | [Vic/Fam] | GCTTGTGGAGAGTGCTGCACAC[A/G]VGTTGGTCTCATCTCTGAGGC |
| *TFGb1* | rs2241713 | [Vic/Fam] | TATAATCACAACCCTGTGAGGTAA[C/G]TGTATTATCTCCATTTGACG |
| *TNF* | rs2256974 | [Vic/Fam] | GCTGGAGAAGGAATGGGCTTC[A/G]TAACCTTGAGCCCTCTTCCCTG |
| *TNF* | rs909253 | [Vic/Fam] | TCACATTCTCTGTTTCTGCCATG[A/G]TTCCTCTCTGTTCCCTTCCTGT |
| *VEGFR* | rs7993418 | [Vic/Fam] | CATGAACTTGAAAGCATTTAC[A/G]TATCTAATGAAGAAACAGAAAGA |
