## Supplementary Table 3 for "Suitability of Machine Learning for Atrophy and Fibrosis Development in Neovascular Age-Related Macular Degeneration"

**Supplementary Table 3.** Minor Allele Frequency differences between Atrophy/non-Atrophy and Fibrosis/non-Fibrosis patients at 36 months.

| | | | Total | Atrophy | | | | Fibrosis | | | |
|---|---|---|---|---|---|---|---|---|---|---|---|
| Gene | SNP | Minor Allele | alleles (n) MAF frec. (%) | Non-Atrophy 36 alleles (n) MAF frec. (%) | Atrophy 36 alleles (n) MAF frec. (%) | allele. Uncor. p-value | allele cor. p | No Fibrosis. 36 alleles (n) MAF frec. (%) | Fibrosis. 36 alleles (n) MAF frec. (%) | Alelle Uncor. p-value | Alelle cor. p |
| *ARMS2* | rs10490924 | T | 281/407<br>40.9 | 186/278<br>40.1 | 91/123<br>42.5 | 0.30 | >1 | 164/250<br>39.6 | 113/151<br>42.8 | 0.42 | >1 |
| *CFB* | rs12614 | T | 96/588<br>14.0 | 67/397<br>14.4 | 29/181<br>13.8 | 0.90 | >1 | 60/352<br>14.6 | 36/226<br>13.8 | 0.82 | >1 |
| *CFB* | rs641153 | A | 47/637<br>6.8 | 34/430<br>7.3 | 12/198<br>5.7 | 0.51 | >1 | 26/386<br>6.3 | 20/242<br>7.6 | 0.53 | >1 |
| *CFH* | rs1061170 | T | 330/354<br>48.2 | 218/246<br>47.0 | 102/108<br>48.6 | 0.73 | >1 | 199/213<br>48.3 | 121/141<br>46.1 | 0.63 | >1 |
| *CFH* | rs800292 | A | 92/592<br>13.4 | 61/403<br>13.1 | 29/181<br>13.8 | 0.90 | >1 | 57/355<br>13.8 | 33/229<br>12.6 | 0.72 | >1 |
| *CFI** | rs4698775 | G | 193/411<br>32.0 | 125/281<br>30.7 | 65/123<br>34.5 | 0.39 | >1 | 110/262<br>30.0 | 82/140<br>36.9 | **0.039** | **0.56** |
| *CFI* | rs17440077 | G | 202/452<br>30.9 | 130/308<br>29.7 | 72/134<br>34.9 | 0.20 | >1 | 117/277<br>29.7 | 85/165<br>34.0 | 0.25 | >1 |
| *CFI* | rs10033900 | T | 304/376<br>44.7 | 205/253<br>44.8 | 91/123<br>44.3 | 0.6 | >1 | 178/234<br>43.2 | 121/137<br>46.9 | 0.37 | >1 |
| *SERPINF1* | rs12603486 | A | 171/515<br>24.9 | 114/350<br>24.5 | 55/157<br>25.9 | 0.77 | >1 | 104/308<br>25.2 | 65/199<br>24.6 | 0.92 | >1 |
| *SMAD7* | rs7226855 | G | 295/375<br>44.0 | 205/251<br>45.0 | 84/120<br>41.2 | 0.39 | >1 | 176/228<br>43.6 | 113/143<br>44.1 | 0.93 | >1 |
| *TFGβ1* | rs2241713 | G | 248/368<br>40.1 | 167/253<br>39.8 | 77/109<br>41.4 | 0.72 | >1 | 146/234<br>38.4 | 98/128<br>43.4 | 0.23 | >1 |
| *TNFα* | rs2256974 | A | 109/503<br>18.8 | 73/345<br>17.5 | 34/150<br>18.5 | 0.81 | >1 | 70/312<br>18.3 | 37/183<br>16.8 | 0.65 | >1 |
| *TNFα* | rs909253 | G | 170/508<br>25.1 | 119/337<br>26.1 | 48/164<br>22.6 | 0.38 | >1 | 99/311<br>24.1 | 68/190<br>26.4 | 0.58 | >1 |
| *VGFR* | rs7993418 | G | 221/467<br>32.1 | 153/313<br>32.8 | 64/148<br>30.1 | 0.53 | >1 | 139/277<br>33.4 | 78/184<br>29.7 | 0.35 | >1 |
