## Supplementary Table 4 for "Suitability of Machine Learning for Atrophy and Fibrosis Development in Neovascular Age-Related Macular Degeneration"

**Supplementary Table S4. Hyperparameter tuning parameters.** Detailed information about the parameters used for the hyperparameter tuning, for each considered ML model.

| **Model type** | **Parameters** | **Values** |
|---|---|---|
| Random Forest | Number of estimators | 50, 287, 525, 762, 1000 |
| | Max depth | 10, 30, 50 |
| | Max features | 'auto', 'sqrt' |
| | Min samples split | 2, 10 |
| | Min samples leaf | 1, 5 |
| | Bootstrap | True, False |
| Extreme Gradient Boosting | Number of estimators | 50, 287, 525, 762, 1000 |
| | Max depth | 10, 30, 50 |
| | Min child weight | 0.1, 5 |
| | Gamma | 0.5, 1.5 |
| | Subsample | 0.6, 1 |
| | Colsample by tree | 0.6, 1 |
| | Learning rate | 0.0001, 0.1 |
| | Reg alpha | 0.0001, 0.1 |
| | Reg lambda | 0.0001, 0.1 |
| Support Vector Machine | C | 0.1, 0.5, 1 |
| | Gamma | 1, 0.01 |
| | Kernel | 'rbf', 'linear', 'sigmoid' |
| | Class weight | None, 'balanced' |
